# Redefining Injury Severity in Youth Sports: Functional Inability to Walk as a Primary Severity Phenotype (n = 4,829)

**DOI:** 10.64898/2026.02.03.26345517

**Authors:** Shinsuke Sakoda, Kimiaki Kawano

## Abstract

**Objectives:** Determinants of functional injury severity in young athletes remain incompletely understood. Conventional assumptions emphasize contact sport participation and skeletal immaturity as major drivers of severe injury; however, severity definitions based on surgery or time loss are strongly influenced by health-care systems and competition contexts. We investigated factors associated with early functional severity after lower-extremity sports injury using a large clinical cohort.

**Methods:** This cross-sectional observational study analyzed 4,829 lower-extremity sports injury cases in athletes aged ≤22 years who presented to a specialized sports injury clinic between January 2017 and November 2025. Severe injury was defined as inability to walk independently at the initial visit. Multivariable logistic regression analysis was performed to evaluate associations between severe injury and age, sex, sport type, injury mechanism, injury location, and physeal status.

**Results:** Severe injury was most strongly associated with acute injury compared with overuse injury (odds ratio [OR], 31.1; 95% confidence interval [CI], 17.7–54.6; p <.001). Injuries involving the knee and ankle were also significantly associated with severe injury (OR, 2.73; 95% CI, 2.04–3.64; p <.001). Female sex showed a modest independent association (OR, 1.40; 95% CI, 1.06–1.84; p =.017), whereas participation in contact sports, physeal status, and age were not independently associated.

**Conclusion:** Early functional severity after lower-extremity sports injury in young athletes was primarily associated with injury acuity and anatomical involvement of the knee and ankle rather than with sport type or skeletal maturity. These findings challenge conventional exposure-based assumptions and support function-oriented phenotyping for injury surveillance and targeted prevention strategies.

## Introduction

Sports injuries in young athletes have become an important issue in orthopaedic and sports medicine practice, reflecting the increasing number of participants and the rising level of competition. ^1, 2^ Large epidemiological studies and systematic reviews have consistently demonstrated a substantial burden of musculoskeletal injuries in pediatric and adolescent athletes, with clear variations according to age, sex, and sport participation patterns. ^3–5^

Lower-extremity sports injuries are particularly common because they are directly related to sport-specific movements such as running, jumping, landing, and directional changes.6 This predominance of lower-extremity involvement has been confirmed across multiple international surveillance systems and youth tournament cohorts.^7, 8^ When severe, these injuries can lead not only to withdrawal from sport but also to long-term functional impairment and adverse effects on athletic development and growth. ^1, 9^ Therefore, prevention of severe lower-extremity sports injuries represents a key challenge in the clinical care of young athletes. ^2, 10^

Among lower-extremity sports injuries in young athletes, severe injuries such as fractures and ligament injuries can markedly restrict walking ability and physical function, resulting in interruption of athletic participation and, in some cases, long-term functional disability. ^11, 12^ Severe sports injuries are often presumed to occur more frequently in contact sports or in skeletally immature athletes. ^9, 13^ However, many of these assumptions are extrapolated from adult populations or overuse-focused cohorts rather than from comprehensive pediatric clinical datasets, and limited quantitative evidence exists regarding which sport characteristics, age groups, or physical backgrounds are actually associated with an increased risk of severe injury in young athletes. ^13–16^

In addition, the definition of severe injury varies widely across studies. ^14, 17^ Many previous reports have defined severity based on the need for surgery or the duration of time lost from sport, both of which are strongly influenced by health care systems, competition structure, and reporting behavior. ^14, 17^ Traditional surveillance metrics may therefore fail to accurately capture the true functional impact of injury at the individual level. ^18–21^ Despite the fact that functional impairment at initial presentation substantially influences early clinical decision-making and return-to-sport trajectories, few studies have evaluated injury severity based on functional status at the time of first clinical assessment. ^14^

Furthermore, most previous studies on lower-extremity sports injuries have focused on specific sports or isolated anatomical regions. ^9, 22^ Large-scale analyses that simultaneously consider clinically relevant variables—such as sport characteristics, injury mechanism, injury site, age, sex, and physeal status—remain scarce. ^13, 16^ As a result, evidence to comprehensively assess determinants of early functional severity and to prioritize preventive interventions in young athletes remains limited. ^2, 10^

To our knowledge, no prior large-scale clinical study has examined determinants of injury severity using an immediate functional outcome measure assessed at first presentation across all types of lower-extremity sports injuries in youth athletes. Evaluating severity through early functional status may provide a more direct and biologically meaningful perspective than traditional exposure- or treatment-based definitions.

Therefore, the purpose of this study was to investigate factors associated with severe lower-extremity sports injuries in a large clinical cohort of young athletes. By simultaneously evaluating multiple demographic, sport-related, anatomical, and developmental variables, this study aimed to clarify determinants of early functional severity and to identify priorities for targeted preventive strategies in youth sports.

## Materials and Methods

### Study Design

This was a retrospective cross-sectional observational study conducted at a single institution.

### Study Population

Lower-extremity sports injury cases in patients aged 22 years or younger (children, adolescents, and young adults) who presented to a specialized outpatient clinic for sports injuries between January 2017 and November 2025 were eligible for inclusion. Lower-extremity injuries sustained during sports participation or attributable to sports activities were included. Non–sports-related injuries, duplicate visits for the same injury, cases with incomplete medical records, and cases lacking appropriate imaging data were excluded.

A total of 4,829 cases were ultimately included in the analysis.

### Definitions

#### Severe Injury

Severe injury was defined as inability to walk independently, including cases requiring crutches or a wheelchair for ambulation. In acute injuries, inability to walk independently was assessed immediately after injury. In overuse injuries, walking ability was assessed at the time of the first clinical evaluation.

Inability to walk independently was selected as a pragmatic and clinically meaningful indicator of functional injury severity, reflecting immediate load tolerance, pain-related functional limitation, and the integrity of weight-bearing structures. This functional criterion is less influenced by health care system factors, treatment preferences, or competitive context than traditional severity measures such as surgical indication or time loss from sport. Because walking ability is routinely assessed at initial clinical presentation and can be evaluated consistently across injury types and age groups, it provides a reproducible proxy for early functional breakdown following lower-extremity sports injury.

Although acute injury and inability to walk may appear conceptually related, many acute injuries (e.g., minor sprains, contusions) do not result in loss of independent ambulation, whereas some overuse conditions can substantially limit function. Therefore, the relationship between injury mechanism and walking ability cannot be assumed a priori and requires empirical evaluation.

#### Injury Mechanism

Injury mechanism was classified as acute injury when symptoms occurred following a clearly identifiable traumatic event, and as overuse injury when symptoms developed gradually due to repetitive loading.

#### Sport Type

Sport type was categorized as non-contact sports or contact sports, based primarily on the presence or absence of interpersonal physical contact during play.

#### Body Region

Injury location was classified into six anatomical regions: pelvis, thigh, knee, lower leg, ankle, and foot. For analysis, the knee and ankle were grouped together as a single region based on shared weight-bearing and impact-absorbing functions, while the pelvis, thigh, lower leg, and foot were combined into an “other regions” category.

Knee and ankle regions were grouped a priori because both represent primary weight-bearing, shock-absorbing joints during gait and stance and have relatively limited degrees of freedom compared with proximal joints, making them more vulnerable to functional collapse when injured. In addition, preliminary univariable analyses demonstrated similar directional associations with inability to walk independently, supporting this grouping for multivariable modeling.

#### Physeal Status

Physeal status was evaluated using plain radiographs and classified as open or closed.

#### Data Collection

Data on age, sex, sport type, injury mechanism, injury location, physeal status, and injury severity were retrospectively collected from medical records and imaging data. Walking ability was routinely documented as a triage assessment item by clinicians at the time of presentation and recorded in the electronic medical record prior to imaging evaluation or definitive diagnosis, thereby minimizing potential classification bias.

#### Statistical Analysis

To identify factors associated with severe injury, a multivariable logistic regression analysis was performed with severe injury (yes/no) as the dependent variable. Independent variables included age, sex, sport type, injury mechanism, injury location, and physeal status.

Results are presented as odds ratios (ORs) with 95% confidence intervals (95% CIs). All statistical analyses were performed using R (R Foundation for Statistical Computing, Vienna, Austria), and statistical significance was set at p < 0.05.

#### Ethics Approval

This study was approved by the institutional review board of the authors’ institution and the requirement for informed consent was waived due to the retrospective study design. The study was conducted in accordance with the Declaration of Helsinki.

## Results

### Baseline Characteristics

A total of 4,829 sports-related lower-extremity injuries in pediatric and adolescent athletes were included in this study (Table 1). The mean age of the patients was 15.0 ± 3.6 years, and 3,532 cases (73.1%) occurred in male athletes.

**Table 1.** Baseline Characteristics of 4,829 Sports-Related Injuries in Youth and Adolescent Athletes.

|  | All cases (n=4,829) |
| --- | --- |
| Age, mean ± SD (years) | 15.0 ± 3.6 |
| Male sex, n (%) | 3,532 (73.1%) |
| Sport type (contact), n (%) | 3,004 (62.2%) |
| Body region |  |
| Pelvis, n (%) | 463 (9.6%) |
| Thigh, n(%) | 397 (8.2%) |
| Knee, n (%) | 1,580 (32.7%) |
| Lower leg, n (%) | 347 (7.2%) |
| Ankle, n (%) | 1,183 (24.5%) |
| Foot, n (%) | 858 (17.3%) |
| Acute onset injury, n (%) | 2,279 (47.2%) |
| Severe injury, n (%) | 376 (7.8%) |

Contact sports accounted for 3,004 cases (62.2%). The most common injury location was the knee (1,580 cases, 32.7%), followed by the ankle (1,183 cases, 24.5%), foot (858 cases, 17.3%), pelvis (463 cases, 9.6%), thigh (397 cases, 8.2%), and lower leg (347 cases, 7.2%).

Acute injuries accounted for 2,279 cases (47.2%). Severe injury, defined as inability to walk independently, was identified in 376 cases (7.8%).

### Factors Associated with Severe Injury

The results of the multivariable logistic regression analysis examining factors associated with severe injury are presented in Table 2 and Figure 1.

**Table 2.** Factors independently associated with inability to walk at presentation in youth and adolescent athletes: Multivariable logistic regression analysis.

| Variables | OR | 95% CI | p-value |
| --- | --- | --- | --- |
| Acute (vs overuse) | 31.1 | 17.7-54.6 | < 0.001 |
| Knee and ankle (vs other parts) | 2.73 | 2.04-3.64 | < 0.001 |
| Female (vs male) | 1.40 | 1.06-1.84 | 0.017 |
| Contact sport (vs non-contact) | 0.95 | 0.73-1.24 | 0.730 |
| Closed physis (vs Open) | 1.01 | 0.68-1.52 | 0.960 |
| Age (per 1-year increase) | 1.05 | 0.99-1.10 | 0.091 |

**Figure 1.**
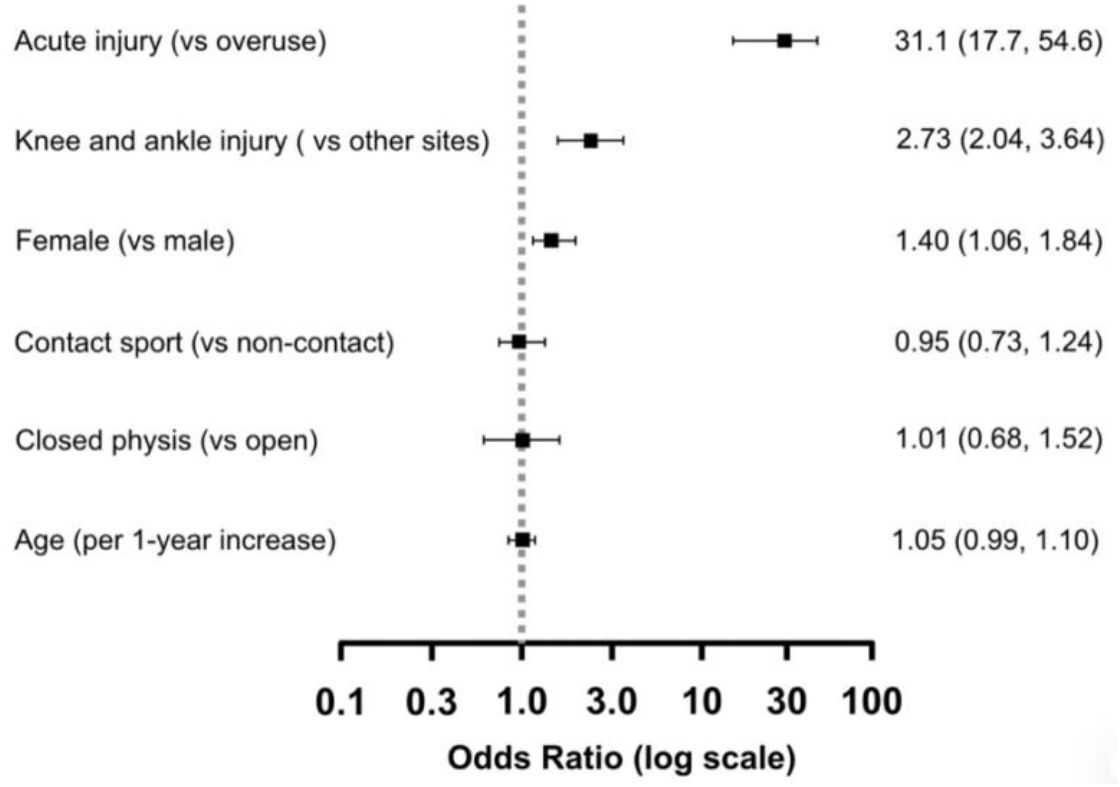
Multivariable logistic regression analysis identifying factors associated with severe lower-extremity sports injury resulting in inability to walk independently in young athletes. Adjusted odds ratios with 95% confidence intervals are shown. Acute injury and knee or ankle injury were strongly associated with severe injury. Female sex showed a modest association, whereas contact sport participation, physeal status, and age were not independently associated.

Acute injury (vs overuse injury) showed the strongest association with severe injury (odds ratio [OR], 31.1; 95% confidence interval [CI], 17.7–54.6; p <.001). Regarding injury location, injuries to the knee and ankle (vs other regions) were significantly associated with severe injury (OR, 2.73; 95% CI, 2.04–3.64; p <.001). Female sex was associated with a higher odds of severe injury compared with male sex (OR, 1.40; 95% CI, 1.06–1.84; p =.017).

In contrast, participation in contact sports (vs non-contact sports) was not significantly associated with severe injury (OR, 0.95; 95% CI, 0.73–1.24; p =.730). Similarly, physeal status (closed vs open) (OR, 1.01; 95% CI, 0.68–1.52; p =.960) and age (per 1-year increase) (OR, 1.05; 95% CI, 0.99–1.10; p =.091) were not significantly associated with severe injury.

## Discussion

In this study, we investigated factors associated with severe lower-extremity sports injuries resulting in inability to walk independently in a cohort of 4,829 young athletes aged 22 years or younger. Severe injury was strongly associated with acute injury and injuries involving the knee and ankle, and the risk was modestly higher in female athletes, whereas participation in contact sports and physeal status were not independently associated with early functional severity.

Traditional definitions of injury severity in sports medicine commonly rely on time loss, surgical indication, or diagnostic labeling.^14–16, 19^ Although these metrics are useful for surveillance and administrative benchmarking, they are strongly influenced by health-care systems, access to specialty care, competitive schedules, and cultural expectations regarding return to play. ^2, 8, 15^ As a result, identical structural injuries may be classified as “severe” or “non-severe” depending on contextual rather than biological factors. ^14–16^ In contrast, inability to ambulate independently at initial presentation represents a pragmatic and clinically actionable marker of early functional collapse that directly reflects the athlete’s capacity to tolerate and compensate for mechanical loading during basic weight-bearing tasks. ^17, 18, 23, 34^

From a clinical perspective, this outcome captures a convergent endpoint of tissue disruption, pain-mediated neuromuscular inhibition, protective motor strategies, and transient loss of load-sharing capacity across the kinetic chain. ^12, 24^–^27, 31^ Importantly, this functional phenotype is immediately observable at first contact, requires no advanced imaging or prolonged follow-up, and aligns naturally with triage decisions, early immobilization strategies, and prioritization of diagnostic resources. ^14, 15, 33^ By focusing on early functional performance rather than downstream care utilization or delayed recovery metrics, the present study shifts the conceptual framework of injury severity toward an exposure-independent, biomechanically grounded construct. ^14–16, 19^ This perspective may facilitate more standardized comparisons across settings and may better reflect the acute physiological consequences of injury that are most relevant to early clinical decision-making. ^17, 18, 34^

The exceptionally large odds ratio observed for acute injury underscores the dominant influence of abrupt mechanical loading on immediate functional failure. ^21, 34^ Acute injuries are characterized by rapid energy transfer that exceeds local tissue tolerance and overwhelms short-term compensatory mechanisms, resulting in sudden loss of load-bearing capacity. ^21, 34^ In contrast, overuse injuries often evolve gradually, allowing partial neuromuscular adaptation, redistribution of mechanical demand, and preservation of basic ambulation despite ongoing structural pathology. ^10, 33^

This distinction suggests that injury mechanism may function less as a direct causal determinant of long-term tissue damage and more as a surrogate marker of the temporal dynamics of mechanical overload and compensation failure. ^17, 34^ Acute traumatic events simultaneously impose peak stresses on musculoskeletal structures and provoke immediate nociceptive input, reflexive muscle inhibition, and protective guarding, thereby amplifying functional impairment beyond the magnitude of isolated structural damage. ^12, 24^–^27, 31^ Conversely, chronic overload permits incremental accommodation and task modification, delaying overt functional collapse even when tissue degeneration is substantial. ^10, 33^

Conceptually, early functional severity may therefore represent a transient “biomechanical shock” state, in which the neuromuscular system temporarily loses its capacity to stabilize and distribute load efficiently across the kinetic chain. ^17, 18, 23, 23^ Recovery of ambulation may occur rapidly as pain, inhibition, and guarding resolve, even though structural healing lags behind. ^12, 24–27^ This dissociation highlights why early functional severity should not be conflated with long-term prognosis, yet remains clinically valuable for acute risk stratification, resource allocation, and immediate management decisions. ^14–16, 19^ Future studies integrating objective measures of loading exposure, neuromuscular activation, and short-term recovery trajectories may further clarify the mechanistic pathways linking acute mechanical insult to early functional collapse. ^17, 34^

The strong association between injuries involving the knee and ankle and inability to ambulate independently likely reflects the central role of these joints in closed-chain load transmission and dynamic stability during stance and gait. ^18, 23, 34^ Both joints serve as primary interfaces between ground reaction forces and proximal segments, regulating shock absorption, propulsion, and postural control. ^23^,^34^ Even modest impairment at these levels can disrupt the integrity of the entire kinetic chain, disproportionately compromising weight-bearing capacity compared with injuries at more proximal or distal segments. ^18, 23^

From a biomechanical standpoint, the ankle modulates initial contact mechanics and mediates rapid adjustments to surface variability, whereas the knee governs sagittal-plane load sharing and energy absorption during mid-stance and deceleration. ^20–22, 23^ Injury-related pain, effusion, or instability at either joint can provoke protective unloading strategies that exceed compensatory reserves, particularly in young athletes with high movement demands and limited tolerance for altered motor patterns. ^12, 24^–^27, 31^ The resulting instability or antalgic gait may render independent ambulation temporarily unsafe or impractical, even when gross structural disruption is not catastrophic. ^22, 23^

Clinically, this finding reinforces the importance of early functional assessment and targeted protection strategies for knee and ankle injuries, regardless of sport category or apparent injury mechanism. ^14–16^ Immediate stabilization, temporary load modification, and early neuromuscular reactivation may mitigate secondary functional deterioration and facilitate safer progression toward rehabilitation. ^28–30, 32, 33^ These results also suggest that anatomical region may serve as a more clinically meaningful axis for early severity stratification than traditional sport-based risk classifications. ^14, 19^

Female sex demonstrated a modest independent association with early functional severity. Although the present dataset does not permit direct evaluation of underlying mechanisms, several plausible contributors warrant consideration. Prior literature has documented sex-related differences in joint laxity, neuromuscular control strategies, muscle strength distribution, and pain perception, all of which may influence acute load tolerance and compensatory capacity during injury. ^11, 12, 24–27, 31^ Subtle differences in early protective motor responses or pain-mediated inhibition may amplify transient functional impairment even when structural injury patterns are comparable. ^12, 24–27^

Importantly, this association should not be interpreted deterministically or as evidence of inherent vulnerability. Unmeasured confounders, including sport-specific exposure profiles, conditioning status, anthropometrics, and access to early supportive care, may partially explain the observed effect. ^17, 18^ Nonetheless, the consistency of this finding across multiple injury contexts suggests that sex-specific neuromuscular and behavioral factors may modulate early functional response to injury and deserve further investigation. ^11, 25, 26^ Incorporating objective biomechanical assessment and early functional performance metrics into future studies may help disentangle biological from contextual contributors. ^12, 31^

In contrast, neither participation in contact sports nor physeal maturity demonstrated a significant association with severe injury. These findings challenge the conventional assumption that contact sports inherently confer higher severity risk and that skeletally immature athletes are intrinsically more vulnerable to severe injury. ^9, 10, 14^ Prior pediatric epidemiological studies have likewise demonstrated substantial heterogeneity of injury risk within sport categories and age groups, highlighting the limitations of broad exposure proxies for meaningful individual risk stratification. ^4, 5^ From a preventive and surveillance perspective, injury acuity and anatomical load-bearing vulnerability appear to represent more proximal determinants of early functional breakdown than sport classification or skeletal maturity. This supports a shift toward function-oriented severity phenotyping rather than reliance on indirect exposure-based classifications alone. ^15, 18^

Among skeletally immature athletes, the relatively high plasticity of bone and soft tissues, combined with lower absolute body mass and variable external loading, may permit injury occurrence without immediate functional collapse severe enough to prevent independent ambulation. ^9^ This may partially explain the absence of a significant association between physeal immaturity and early functional severity in the present cohort.

From a clinical and preventive perspective, these findings indicate that preventive strategies should prioritize knee and ankle injury mechanisms, particularly in female athletes, rather than applying uniform risk assessments based solely on age or sport participation. Indeed, randomized and controlled trials have demonstrated that structured neuromuscular and warm-up–based prevention programs can substantially reduce the incidence of serious knee and lower-extremity injuries in youth populations. ^22–24, 28^–^31, 33, 34^ In addition to neuromuscular conditioning, targeted instruction in landing and cutting techniques, as well as assessment and intervention addressing hip mobility and foot function, may further reduce excessive focal loading and mitigate early functional collapse. ^11, 12, 19, 24^

### Clinical Perspective

Functional severity at presentation, defined by inability to walk independently, reflects early biomechanical and load-bearing failure more directly than traditional exposure-based severity definitions based on time loss or surgical intervention. ^15, 18^ Acute injury and knee or ankle involvement represent the primary drivers of early functional collapse, whereas sport type and skeletal maturity do not independently determine severity.

These findings support a shift toward function-oriented severity phenotyping in early clinical triage, enabling clinicians to prioritize timely protection, diagnostic evaluation, and targeted rehabilitation strategies based on immediate functional impairment rather than indirect exposure proxies. ^14–16, 19^ In particular, heightened vigilance and early load-modifying interventions may be warranted for acute knee and ankle injuries, especially in female athletes. ^17, 18^

Integration of structured neuromuscular prevention programs, movement quality assessment, and progressive return-to-loading strategies may further mitigate early functional breakdown and support safer long-term sport participation in youth populations. ^22–24, 28–34^

### Limitations

This study has several limitations. First, it was a single-center retrospective study, and detailed external factors such as competition level and injury circumstances could not be fully evaluated. Second, although inability to walk independently is a clinically clear outcome, it does not directly assess long-term prognosis or return-to-sport outcomes. In addition, functional walking ability at presentation does not directly capture imaging-based structural severity, surgical necessity, or long-term recovery trajectories. Although this measure reflects early functional impact, future longitudinal studies integrating structural injury grading and recovery trajectories are warranted to further validate its prognostic relevance. Future studies should further clarify the specific injury mechanisms within acute injuries that contribute to greater severity, as well as the mechanisms underlying sex-related differences in knee and ankle injuries.

Moreover, the single-center nature of this cohort may limit the generalizability of the findings to other health-care systems, competitive environments, and referral patterns. Because the study population consisted of athletes presenting to a specialized sports injury clinic, milder injuries managed in primary care or community settings may have been underrepresented, potentially inflating the observed proportion of functionally severe presentations. Because walking ability was recorded before diagnostic clarification, this measure reflects true functional status at presentation rather than being influenced by knowledge of injury severity.

Additionally, although inability to walk independently provides a pragmatic and clinically meaningful indicator of early functional severity, this measure may be influenced by individual pain tolerance, psychological responses to injury, and short-term protective behavior, which were not systematically captured in the present dataset. The outcome reflects acute functional status rather than structural tissue damage, surgical necessity, or long-term recovery trajectory, and should therefore be interpreted as a marker of early functional impact rather than definitive injury prognosis.

Finally, several potentially relevant confounders—including body mass index, sport-specific exposure intensity, competitive level, prior injury history, neuromuscular conditioning status, and psychosocial factors—were not available for adjustment. Future multicenter and prospective studies incorporating objective exposure metrics, biomechanical assessment, and longitudinal outcomes will be necessary to validate the external applicability of the present findings and to refine clinically actionable severity stratification models.

## Conclusion

Acute injuries were associated with a markedly higher risk of severe lower-extremity sports injury than overuse injuries in young athletes aged 22 years or younger. Female athletes and injuries to the knee and ankle were also associated with greater injury severity, whereas participation in contact sports and physeal maturity were not. These findings suggest that preventive strategies should prioritize knee and ankle injuries, particularly in female athletes, rather than relying on uniform risk assessments based solely on sport type or age. Accordingly, early clinical attention and targeted preventive strategies should be prioritized for acute knee and ankle injuries in young athletes.

## Acknowledgements

The authors would like to thank Erina Murata for her valuable assistance with data collection and organization in the Department of Sports Medicine.

## Data Availability

The data that support the findings of this study are available from the corresponding author upon reasonable request.

## Biographical Notes

Shinsuke Sakoda, MD is an orthopaedic surgeon and sports medicine physician at the Department of Sports Medicine, Ashiya Central Hospital, Fukuoka, Japan. His clinical and research interests focus on youth sports injury epidemiology, lower-extremity biomechanics, injury prevention, and return-to-sport strategies.

Kimiaki Kawano, MD is an orthopaedic surgeon at the Department of Orthopaedic Surgery, Ashiya Central Hospital, Fukuoka, Japan, with clinical interests in sports-related musculoskeletal injuries and surgical treatment of athletic injuries.

